# How should we study the indirect effects of antimicrobial treatment strategies? A causal perspective

**DOI:** 10.1101/2025.03.28.25324855

**Authors:** Juan Gago, Christopher Boyer, Marc Lipsitch

## Abstract

Effective antimicrobial stewardship requires unbiased assessment of the benefits and harms of different treatment strategies, considering both immediate patient outcomes and patterns of antimicrobial resistance. In principle, these benefits and harms can be expressed as causal contrasts between treatment strategies and therefore should be ideally suited for study under the potential outcomes framework. However, causal inference in this setting is complicated by interference between individuals (or units) due to the indirect effects of antibiotic treatment, including the spread of resistant bacteria to others. These indirect effects complicate the assessment of trade-offs as benefits are mostly due to the direct effects among those treated, while harms are more diffuse and therefore harder to measure. While causal frameworks and study designs that accommodate interference have previously been proposed, they have been applied predominately to the study of vaccines, which differ from antimicrobial interventions in fundamental ways. In this article, we review these existing approaches and propose alternative adaptations tailored to the study of antimicrobial treatment strategies.

## 1. Introduction

Antimicrobial resistance (AMR) is a complex and growing threat to global health. It increases patients’ risk of serious illness and mortality and complicates clinicians’ choice of antimicrobial treatments. Globally, AMR threatens the effectiveness of antimicrobials, the therapeutic cornerstone of modern infectious disease management. Furthermore, AMR disproportionately affects vulnerable populations, such as immunosuppressed patients [1], magnifying the clinical and public health burden. Effective interventions are necessary to limit resistance while continuing to maximize the benefits of antimicrobial therapy [2].

To evaluate the harms and benefits of interventions to curb AMR, we need a framework to formalize the causal effects of antimicrobial treatments on microbiological and clinical outcomes and study designs that can be used to empirically estimate these effects. Causal inference methods have become essential tools for formalizing treatment effects. However, their application to AMR, where treatment and broader effects due to transmission exist, has been limited.

A major methodological challenge in developing a causal framework for AMR, common across infectious diseases, is the presence of *interference* —when one person’s treatment affects others’ outcomes, violating standard assumptions [3]. Indirect effects are especially important in AMR. Ignoring them can lead to incomplete assessments of an intervention’s benefits and harms by focusing solely on individual outcomes and overlooking how treatment policies influence population-level infection and resistance patterns. Many resistance-related interventions are justified precisely by their effects on others, such as isolating carriers, [4], treatment as prevention [5], and antimicrobial cycling [6]. Though often studied with transmission-dynamic models, a formal causal framework for these strategies does not yet exist.

Over the past three decades, methods for causal inference under interference have been developed [7–10], mainly for vaccination. However, vaccines differ substantially from antimicrobial and AMR interventions: they’re typically assigned pre-exposure to prevent infection, while antimicrobials are often given post-exposure, to symptomatic individuals, with treatments updated based on diagnostic data. There is no widely adopted causal framework for estimating indirect and overall effects of antimicrobial treatments.

In this paper, we address this gap by extending causal inference methods to antimicrobial treatments and AMR. We define direct and indirect effects in this context, considering that the effects of interest can be different measures of clinical success (e.g., microbiological clearance) as well as adverse outcomes, such as treatment failure and development of AMR infections. We propose modifications to study designs —such as the two-stage randomization used in vaccine studies— to capture the indirect effects of antibiotics. Additionally, we highlight the benefits of estimating well-defined per-exposure effects to obtain accurate estimates of direct effects. Using directed acyclic graphs (DAGs) and single-world intervention graphs (SWIGs), we analyze both direct and indirect effects of antimicrobial treatment strategies and discuss specific challenges that researchers might encounter. The insights from this framework are valuable for designing and evaluating interventions and policies aimed at controlling the spread of AMR and optimizing antibiotic use.

## 2. Motivating example: empiric treatment of bloodstream infections

Vancomycin is commonly used empirically (before pathogen identification) for bloodstream infections (BSI) to cover *Staphylococcus aureus* (including MRSA). However, its use can promote the growth and spread of vancomycin-resistant *Enterococcus* (VRE) by suppressing susceptible *enterococci* and altering gut microbiota [11]. In contrast, daptomycin is a last-resort antibiotic effective against both MRSA and VRE, usually reserved for resistant infections [12]. Thus, there are trade-offs in using daptomycin versus vancomycin as empiric therapy, especially in settings with high MRSA/VRE prevalence. Let’s consider a hypothetical trial comparing vancomycin versus daptomycin as empiric treatment in an ICU for suspected BSI. Traditionally, this would be evaluated in a randomized trial, often set up as a non-inferiority trial: patients with suspected BSI are randomly assigned to one of the arms, vancomycin or daptomycin. Outcomes could include microbiological clearance of infection, clinical resolution, patient mortality, and the development of antimicrobial resistance, specifically an infection that yields a subsequent positive culture for vancomycin-resistant Enterococci (VRE). Another outcome might be incident VRE colonization, detectable if baseline screening is available. Typically, trials measure outcomes only in the treated patients. However, treatments can also have indirect effects on patients’ contacts, fueling the spread of resistance. For instance, vancomycin in a patient may promote VRE growth in that patient’s bowel by eliminating susceptible competitors (vancomycin-susceptible enterococci). The patient then sheds VRE, facilitating transmission to others, increasing the the risk that contacts develop VRE infection [13]. These indirect effects are particularly salient in settings like hospitals or nursing homes (where healthcare-associated infections are common). This scenario illustrates interference: vancomycin administered to one patient can promote resistant strains that spread and undermine the efficacy of treatment in others. Notably, if trial inclusion requires suspected BSI, these secondary cases might not become participants (e.g., if transmitted VRE causes a UTI instead of BSI). Whether they enroll or not, a VRE infection caused by another patient’s treatment would not be accounted for in trial outcomes. Quantifying these indirect effects is crucial to capture the full range of risks and benefits of each strategy. However, such effects are difficult to study in a conventional individually randomized trial. Similar challenges arise in observational studies that seek to emulate this comparison.

## 3. Defining direct and indirect causal effects under interference

To begin fixing concepts, consider the simple scenario in which there are just two possible individuals interacting in a hospital setting (e.g. sequential patients admitted to an isolated ICU room) indexed by *i* and *j*. At admission, we define the following variables: the individual’s true infection status as *I*_*i*0_, *I*_*j*0_; whether infection is suspected (e.g. presenting symptoms) as *S*_*i*0_, *S*_*j*0_. A suspected infection can trigger an initial blood culture (*Y*_*i*0_, *Y*_*j*0_) which, for simplicity, we assume is measured without error. The possible values of both infection status and culture result are no infection/negative culture, susceptible infection/culture, and resistant infection/culture.For example, *Y*_*it*_ for individual *i* at time *t* is defined as

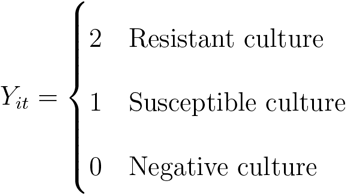

Let (*A*_*i*0_, *A*_*j*0_) denote the empiric antimicrobial treatment of person *i* and *j* prior to pathogen identification and (*A*_*i*1_, *A*_*j*1_) denote the concordant treatment after the results of the culture are obtained (time 1). Following our example, assume that there are two possible antibiotic treatments, one for which resistant species of bacteria exist (vancomycin) and another for which resistance is uncommon (daptomycin). Therefore, define for indiviudal *i* at time *t*

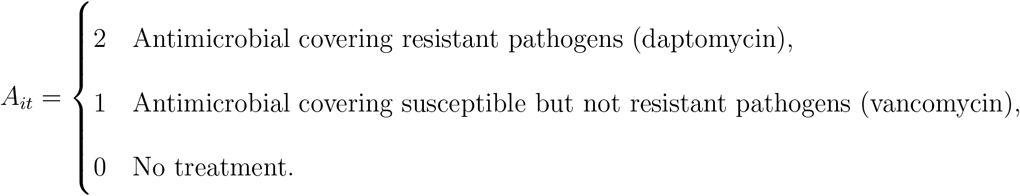

At a future follow up —say 7 days after admission— we define final infection status (*I*_*i*7_, *I*_*j*7_) and culture result (*Y*_*i*7_, *Y*_*j*7_).We also define an intermediate microbiome result (*M*_*i*1_, *M*_*j*1_) to reflect the possibility of carriage (but not infection). The assumed causal relationship between these variables is captured in the causal directed acyclic graph (DAG) of Figure 1. As in our motivating example, consider the causal effect of empiric treatment. The potential outcomes framework defines causal effects in terms of contrasts of hypothetical interventions, which may be contrary to fact. Let 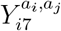 denote the potential culture status of person *i* if the empiric treatment status of person *i* and person *j* were set to (*a*_*i*_, *a*_*j*_). Thus for example 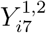 is the potential outcome of person *i* at the 7-day follow up if person *i* receives empiric vancomycin while person *j* receives empiric daptomycin; and 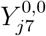 denotes the potential outcome of person *j* if both person *i* and person *j* are untreated. In general, each individual has 9 potential outcomes: 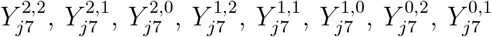, and 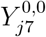. However, we focus on the case that person *i* is admitted prior to person *j*, in which case transmission can only occur from person *i* to person *j*. Therefore, person *i* is unaffected by the treatment of person *j* implying 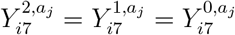 for all *a*_*j*_ and thus we can simplify to only 3 potential outcomes for person 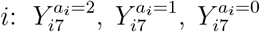. Similarly, because initial culture result for person *j* is unaffected by their empiric treatment, assuming initial cultures are always taken prior to initiation of treatment, we have 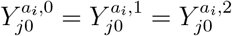 for all *a*_*i*_ and we can again simplify to 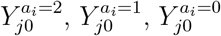.

**Figure 1.**
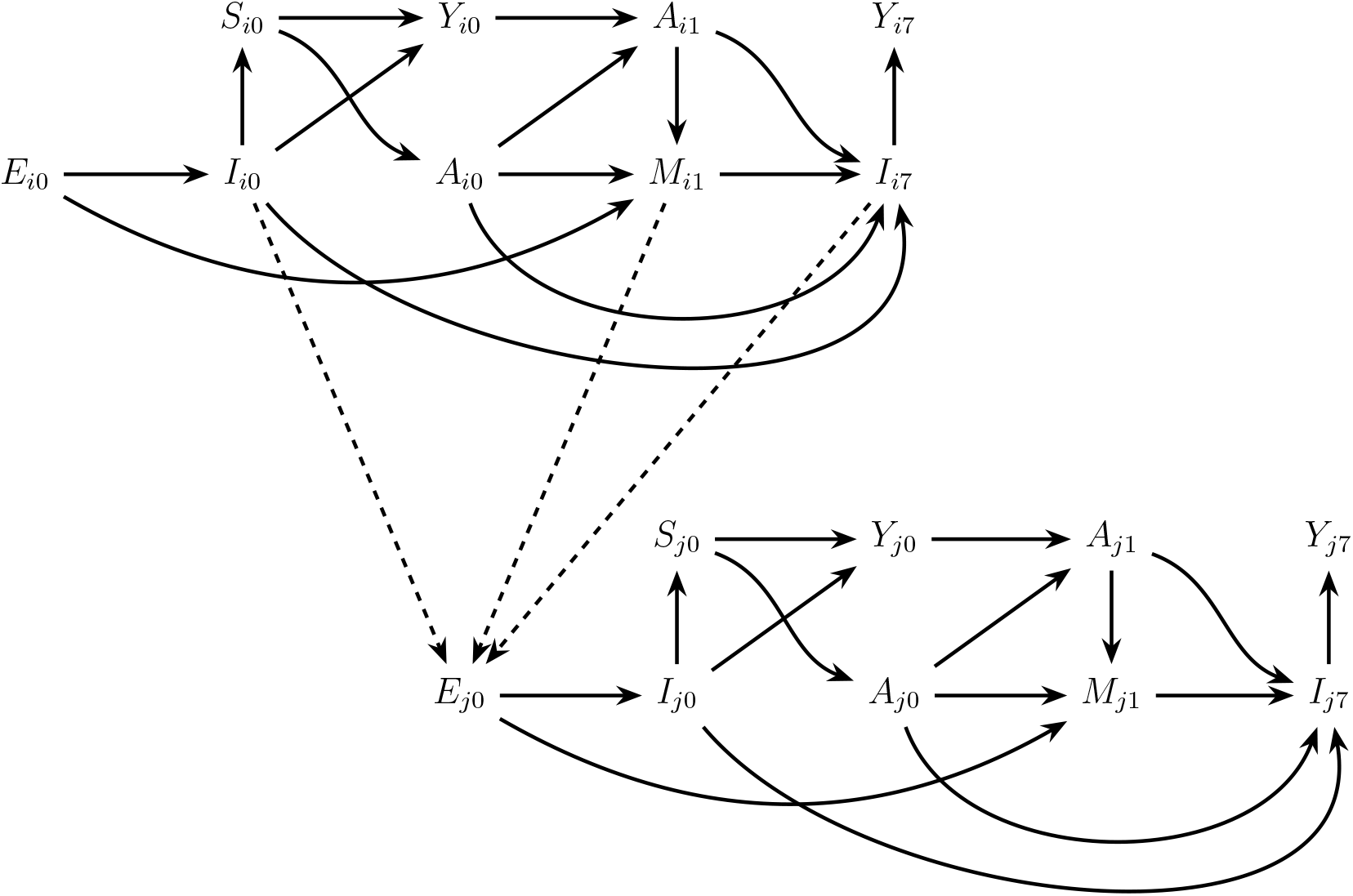
Causal DAGs showing possible interference mechanisms for antimicrobial treatment among two individuals indexed by *i* and *j*. Each subject *i* (top) and *j* (bottom) has cultures for at least two time points: 0 and 7. The node *E*_*i*0_ represents an *exposure* to a pathogen at baseline, which can lead to an *infection I*_*i*0_ suspected by the presence of *symptoms* denoted by *S*_*i*0_. This prompts an *empiric treatment A*_*i*0_, and the initiation of microbiological laboratory tests, which provide an *initial culture result Y*_*i*0_. Guided by *Y*_*i*0_, the clinician may adjust the treatment, yielding *A*_*i*1_. Both *A*_*i*0_ and *A*_*i*1_ can alter the *microbiome M*_*i*1_, which, in turn, can influence the final culture result *Y*_*i*7_ at follow-up. Notably, there is *interference* across subjects, as one subject’s infection or microbiome changes can affect another’s exposure. Specifically, *I*_*i*0_, *I*_*i*7_ or *M*_*i*_ may alter *E*_*j*0_ if subject *i* transmits the pathogen to subject *j*. These dashed cross-subject arrows capture real-world interference pathways in which clinical interventions for one person may have downstream effects on the exposures and infection risks of others.

Notice, under this notation, the potential outcomes of person *j* depend on the treatment status of person *i*. This is referred to as *interference* and is contrary to most introductory literature in causal inference where it is assumed that a person’s potential outcome may be defined independently of the treatment status of others. However, this assumption is implausible for an infectious disease because pathogens are generally transmitted from one individual to another. Returning to our motivating example, comparing effects of vancomycin vs. daptomycin, consider the following plausible scenario: when person *i* is treated with vancomycin it can lead to an overgrowth of VRE which they then transmit to person *j*. When person *i* is instead treated with daptomycin all *enterococci* may be eliminated and therefore person *j* has no additional risk of infection. This implies that 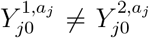 and therefore the no interference assumption is invalid.

We can now formalize the notion of direct and indirect effects of antimicrobial treatments by focusing, in turn, on the outcomes of person *i* and person *j*. To introduce the notion of expectation, we expand consideration to a superpopulation of *K* pairs of admitted patients where interaction is allowed within but not between pairs.

The direct effects of treatment for person *i* may be defined as contrasts between 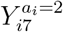 and 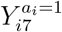, such as

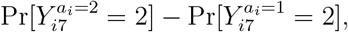

representing resistant infection 7 days after admission when treated empirically with daptomycin versus vancomycin. When combined with their culture result at admission this creates three strata in which we can compare the direct effects of vancomycin vs. daptomycin on person *i* with some of the most sensible contrasts being:

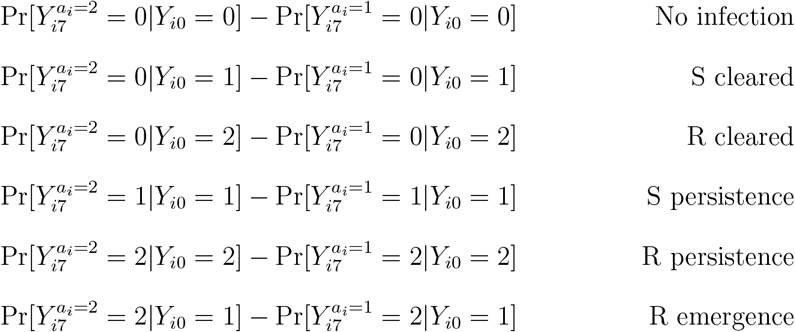

When the presenting infection type is not controlled and the effects of antibiotic choice are heterogeneous across indiviudals its possible that, for instance, the stratum-specific effects above may vary depending on the types of individuals who present with susceptible or resistant infections (discussed further in the next section).

The indirect effects of treatment of person *i* on the outcome of person *j* may be defined as contrast between 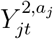 and 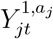, such as

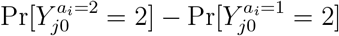

and

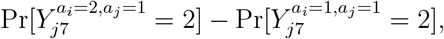

representing resistant infection in person *j* at baseline and follow up when person *i* is treated empirically with daptomycin versus vancomycin and acknowledging that, depending on the mechanism, the treatment of person *i* may affect either the initial (*Y*_*j*0_) or follow up culture result (*Y*_*j*7_) or both.

Both direct and indirect effects are represented graphically via the DAGs in Figure 1. Building on the work of Ogburn and VanderWeele [14], arrows connecting nodes of the same individual (e.g., from *I*_*i*0_ to *A*_*i*0_, or from *A*_*i*0_ to *M*_*i*_) represent *direct effects*. Arrows *between* individuals (e.g., from *I*_*i*0_ or *M*_*i*_ to *E*_*j*0_) capture *indirect effects*—the essence of interference—where one person’s infection or pathogen shedding can expose another person and cause a new infection. Figure 1 represents several possible interference pathways. For example, one route may be that subject *i*’s infection persists or relapses (node *I*_*i*7_), thereby exposing subject *j*. Another possibility is that *A*_*i*0_ or *A*_*i*1_ modifies *M*_*i*_, creating the conditions to allow overgrowth and transmission to *j* of a pathogen. In reality, multiple pathways can operate simultaneously, as indicated by the cross-subject arrows from *i* to *j*. Furthermore, we can extend our graphs to include potential outcomes through single world intervention graphs (SWIGs) as shown in Figure 2. SWIGs represent hypothetical interventions on nodes through a node-splitting transformation [15]. All nodes that are descendants of the split node (intervention node) are then potential outcomes under the specified intervention.

**Figure 2.**
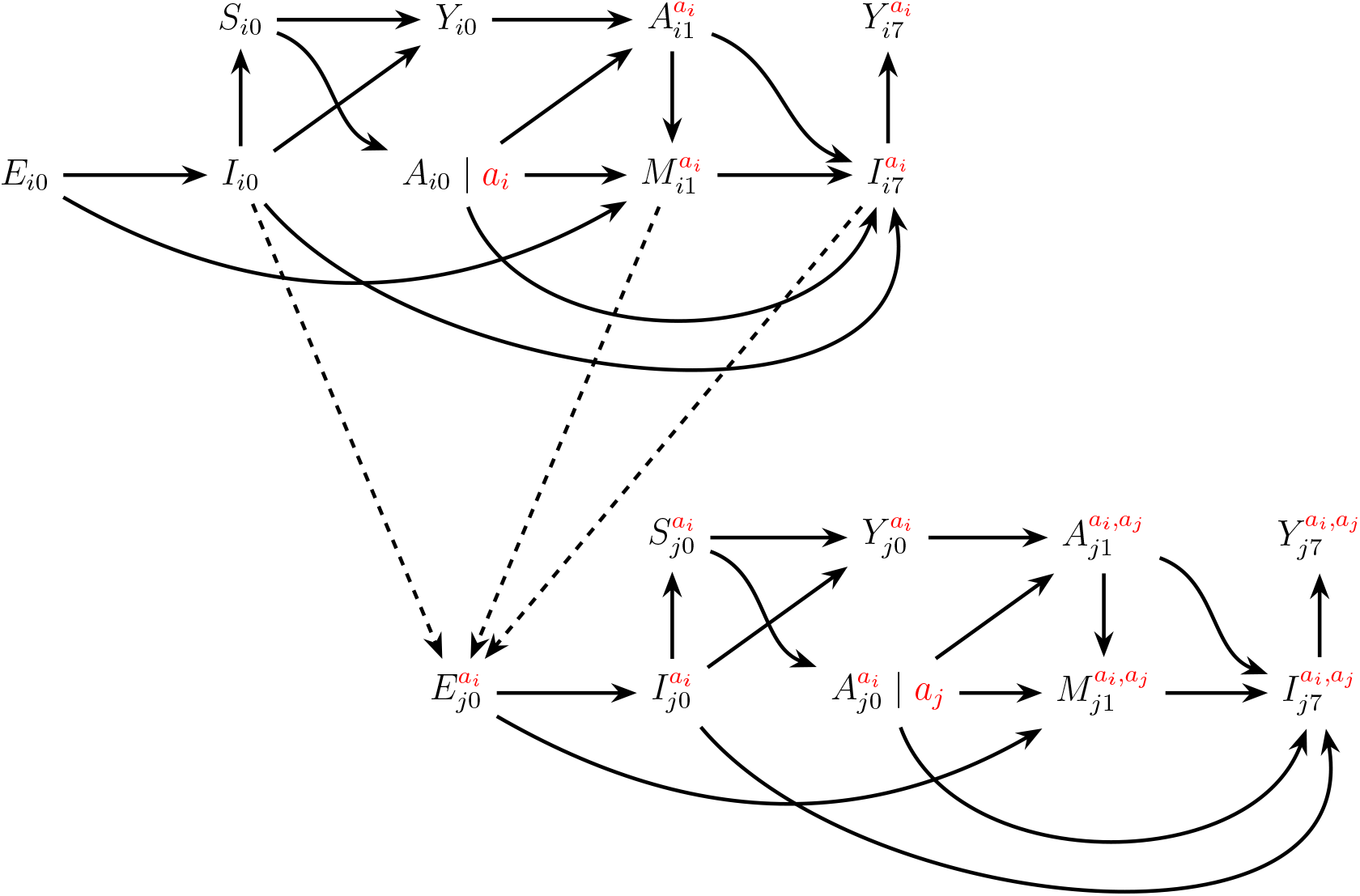
Single world intervention graph (SWIG) showing hypothetical interventions on empiric treatment of individuals *i* and *j*. The influence of the treatment of *i* on *j* is shown explicitly as all *j* nodes are now adorned by *a*_*i*_ indicating the potential outcome of person *j* when person *i* is treated with empiric treatment *a*_*i*_.

## 4. How should we study causal effects of antimicrobial treatments under interference?

Broadly, two approaches exist for studying causal effects under interference in infectious diseases. The first directly manipulates transmission to isolate direct and indirect effects; the second uses clustered groups with two-stage randomization to separate within-vs. between-group effects.

## 5. Approach 1: Isolating direct and indirect effects in individuals via controlled exposure to pathogen

The first approach to interference attempts to isolate direct and indirect effects through controlled exposure, a design know as “challenge trial”. In this experimental design, individuals are placed in isolation and experimentally exposed to both the pathogen of interest and the treatment. It removes the possibility of interference because isolation prevents all indirect effects via transmission from external sources. While the design has been conducted in the past under certain restricted scenarios [16], it is often infeasible due to ethical and logistical considerations.

We can formalize antimicrobial effects under a challenge trial by defining exposure *E*_*i*0_ for individual *i* as:

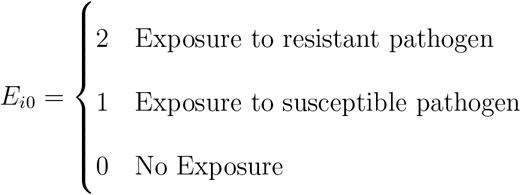

where, in a real challenge trial, the method and dose of each inoculum would be specified. Traditionally, subjects are individually isolated in which case the design targets the so-called “per-exposure effect” [17], i.e. the direct effect of exposure and treatment on the same individual.

### 5.1 Direct effects

We can represent hypothetical challenge interventions—randomizing individuals to different exposures—using a SWIG in which we intervene on exposure. (Figure 3). Contrasts between potential outcomes under these interventions could then be used to quantify the direct harms or benefits for the treated individual (direct effects) of different treatment strategies. For example, we could contrast the probability of eradication at day 7 (*Y*_*i*7_ = 0) when using a treatment option including antimicrobials covering resistant strains (*a*_*i*_ = 2) compared to using antimicrobials covering only susceptible strains (*a*_*i*_ = 1) given an exposure to resistant pathogen (*e*_*i*_ = 2),

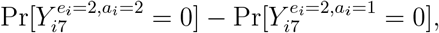

or a susceptible pathogen (*e*_*i*_ = 1)

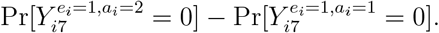

**Figure 3.**
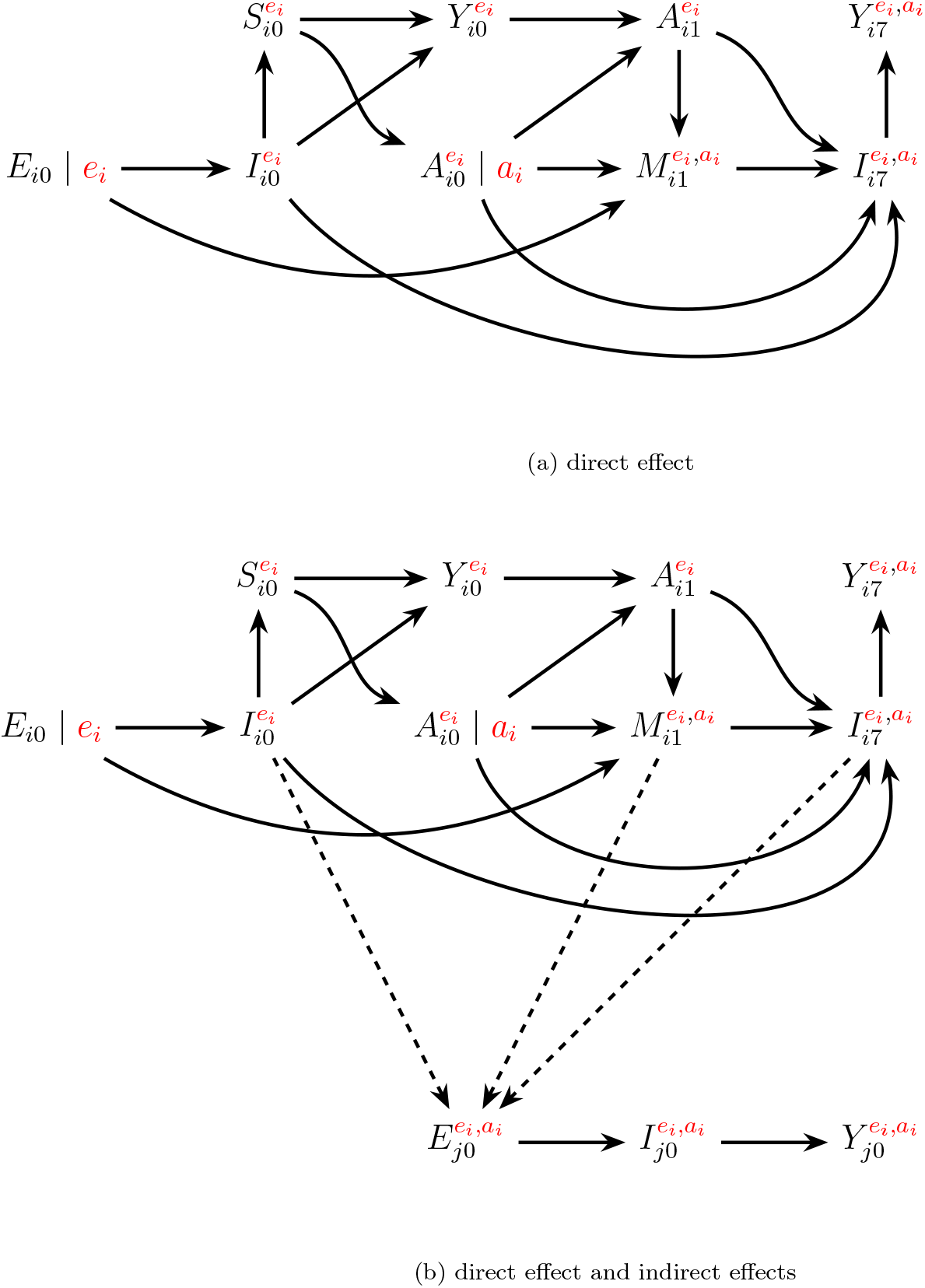
SWIG for possible joint intervention on *E*_*i*0_ and *A*_*i*0_ in a randomized human challenge trial. The top panel shows the case where individual *i* is isolated, receives challenge exposure *E*_*i*0_ = *e*_*i*_, and then is empirically treate d with *A*_*i*0_ = *a*_*i*_. It focuses on direct effects on person *i*. The bottom panel considers an alternative challenge trial in which person *i* and *j* are put in isolation together and *i* receives the challenge exposure while *j* is allowed to interact with them “naturally”.

Controlled exposure allows us to study the possible heterogeneity in response to antibiotic treatment based on many factors, including a unique set of comorbidities or other patient characteristics (e.g., age, pregnancy, etc.). Few study designs directly confirm that results from *in vitro* susceptibility tests translate effectively *in vivo*. Examining the per-exposure effect in a challenge trial like this would be the closest equivalent to estimating, in humans, the direct *in vivo* effect of an antimicrobial on an infection.

### 5.2. Indirect effects

Challenge trials are use to estimate direct effects on the exposed individual. However, it would be possible to measure the amount of susceptible and resistant pathogens an infected individual sheds (and how this is affected by antimicrobial treatment) or it would be possible to experimentally replicate the process of transmission through controlled contact with a second individual [18]. These modifications in the study design would help to estimate potential indirect effects.

In Figure 3(b) we represent the exposure of person *j* when they are placed in contact with person *i* with *E*_*j*_. If, as assumed, this is done in a way that mimics “natural” exposure, i.e. we just place them in the same room, then this second exposure is not “intervened” on as such and, therefore, we do not split the node in the SWIG.

### 5.3. Observational emulation

Challenge trials are rarely conducted in practice due to ethical constraints, as they are justifiable only when the risk of severe infection is extremely low. In the absence of such trials, observational data could be used to emulate them. For instance, one could analyze outcomes in patients with suspected bacterial infections who are effectively isolated with their household members, or in hospitalized patients in single-bed rooms. In both scenarios, estimating the per-exposure effect would require baseline exposure data (e.g., screening cultures to determine exposure status), and since exposure and treatment are not randomly assigned, confounding must be addressed. This implies finding quasi-random variation in exposure or treatment, or measuring a comprehensive set of covariates to adjust for confounding. Special attention is needed to account for common causes of both exposure and outcome, as well as treatment and outcome [17].

Per-exposure effects clarify mechanisms under controlled conditions but do not directly translate to outcomes in populations with natural, uncontrolled exposure. One way to address this limitation is by using these estimates to inform individual-based or mathematical models that simulate infection dynamics under assumed exposure mechanisms.

## 6. Approach 2: Studying population effects under real-world exposures via two-stage randomization

A second approach to studying causal effects under interference is to identify clusters of individuals such that individuals within clusters interact, but those across clusters do not. Direct and indirect effects can be studied through two stages of randomization designs. In the first stage, clusters are randomized to different saturation levels of individual treatments (e.g. treat 30% with vancomycin and 70% with daptomycin). In the second stage, individuals within groups are randomized to receive treatment according to the assigned saturation levels of their group. This design was first described in canonical work by Halloran & Struchiner [3] as applied to vaccines, but it has also been used to study, for instance, the equilibrium effects of economic interventions [7].

In the following sections, we describe possible adaptations of the two-stage design to the study of antimicrobial treatments. A study that would closely resemble the canonical two-stage randomized trial design for vaccines, but applied to antimicrobials, is the evaluation of pre-exposure prophylaxis, since antimicrobials are given prior to exposure, as with vaccines. We briefly discuss this in Appendix A.

### 6.1. Design 1: Post-exposure treatment of suspected infection

As in our motivating example, many antimicrobial treatments target individuals with suspected infections, creating challenges for the canonical designs since only a subset of the population —those with suspicion of infection— is eligible for treatment at any given time. Thus, allocation programs become *dynamic* interventions rather than the *static* interventions in vaccine trials. Furthermore, eligibility evolves over the follow-up period and is affected by treatment in previous time periods because eligibility is based on suspected infection (which is acquired from exposure to previously infected and possibly treated individuals). Finally, post-exposure studies involve complex sampling schemes: we typically only enroll and measure outcomes in those who present as a suspected infection at some point over the follow-up period, we often do not measure outcomes of individuals in the cluster who do not. The latter has implications for the types of indirect effects that we can study.

Following Hudgens & Halloran [8] and VanderWeele & Tchetgen Tchetgen [10], we expand our previous setup as follows. Consider a sample of *K* clusters, indexed by *k* = 1, …, *K*, where now each cluster is comprised of *n*_*k*_ individuals (instead of just two). As before, individuals within clusters interact but there is (approximately) no interaction across clusters. The antimicrobial treatment received by individuals in cluster *k* is represented by the vector (vectors represented in **bold**) 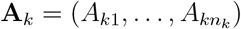, where *A*_*kj*_ is the treatment received by individual *j*. In the MDA setting treatment only occurs at one time point and, for simplicity, we define each *A*_*kj*_ to be binary (0: no treatment, 1: azithromycin). We define **A**_*k,−j*_ as the treatments received by the other individuals in cluster *k* excluding individual *j*. We refer to **A**_*k*_ as the cluster-level treatment or allocation program. The set of all possible treatments for cluster *k* is *𝒜* (*n*_*k*_), meaning **A**_*k*_ can take one of 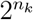 possible values. Let 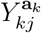 denote the potential outcome of individual *j* in cluster *k* at the end of follow up (e.g. 30 days) under allocation program **a**_*k*_ and let 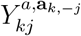 denote the potential outcome when individual *j* has treatment status *a* and the rest of the group *k* has treatment status **a**_*k,−j*_. As before we define *Y*_*kj*_ to be tri-level culture result (0: negative culture, 1: susceptible culture, 2: resistant culture). To ease notation, consider a simple scenario where 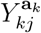 and 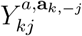only depends on **a**_*k*_ and **a**_*k,−j*_ via the proportion of individuals who are treated which we denote by *π*, i.e. 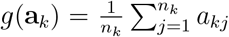. In this case, we can simplify potential outcomes to 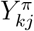 and 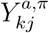. We also allow for 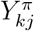 and 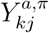 to be random rather than fixed by considering sampling from a superpopulation of clusters.

We enrich the previous setup by indexing the time each variable is measured using *t* = 0, …, *T*. We define an example dynamic post-exposure allocation program which assigns individuals within clusters who present with suspected infection to empiric antimicrobial treatment *A*_*t*_ = 1 (e.g. vancomycin) with probability *π* and empiric treatment *A*_*t*_ = 2 (e.g. daptomycin) with probability 1 *−π*. That is, we assign treatment program **A**_*t*_ in each cluster at time *t* according to the probability mass function,

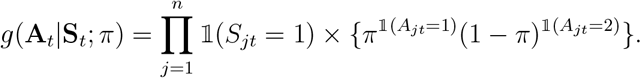

This implies that, at each time point, on average, 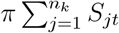 individuals are treated and 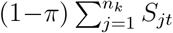 individuals are untreated within each cluster. Taken over time this essentially defines a series of nested trials among those admitted with suspected infection at each time point. Let 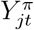 denote the potential outcome of an individual *j* within a cluster at time *t* under an intervention that assigns antimicrobial treatment according to *g*(**A**_*t*_|**S**_*t*_; *π*) and let 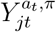 denote the potential outcome of an individual assigned treatment status *A*_*t*_ = *a*_*t*_ at time *t* while the rest of their cluster is assigned treatment according to *g*(**A**_*t,−j*_|**S**_*t,−j*_; *π*). As before we assume potential outcomes only depend on the proportion treated within cluster and are invariant to which specific individuals are treated.

Under this design, we can define a direct effect on resistance at time *t* + 7, i.e. 7 days after admission, as

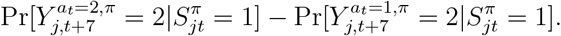

with other direct effects defined based on follow up and initial culture results as in Section 3, e.g. the direct effect on the emergence of resistance is

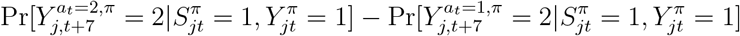

and the head-to-head comparison of daptomycin versus vancomycin on clearance of susceptible baseline infection is

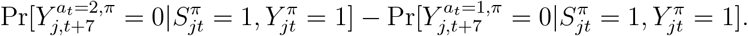

In contrast with the canonical two-stage design, here there are multiple direct effects (one for each time point) each defined for a potentially non-overlapping subgroup composed of those with suspected infection at time *t*. Because exposure is affected by prior antimicrobial treatment assignment in the cluster, in general we would expect the effects to vary as the composition of resistance among those presenting with suspected bacterial infection changes over time. Under randomization, the direct effects on resistance above are identified by

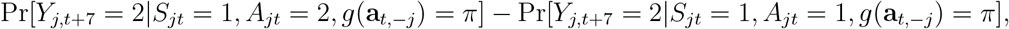

which is the within-cluster comparison of those treated versus untreated after presenting with suspected bacterial infection. The results for direct effect on both AMR emergence and effectiveness in treating the infection are also identified by the within-cluster comparison among their admission culture subgroups.

**Figure A.4:**
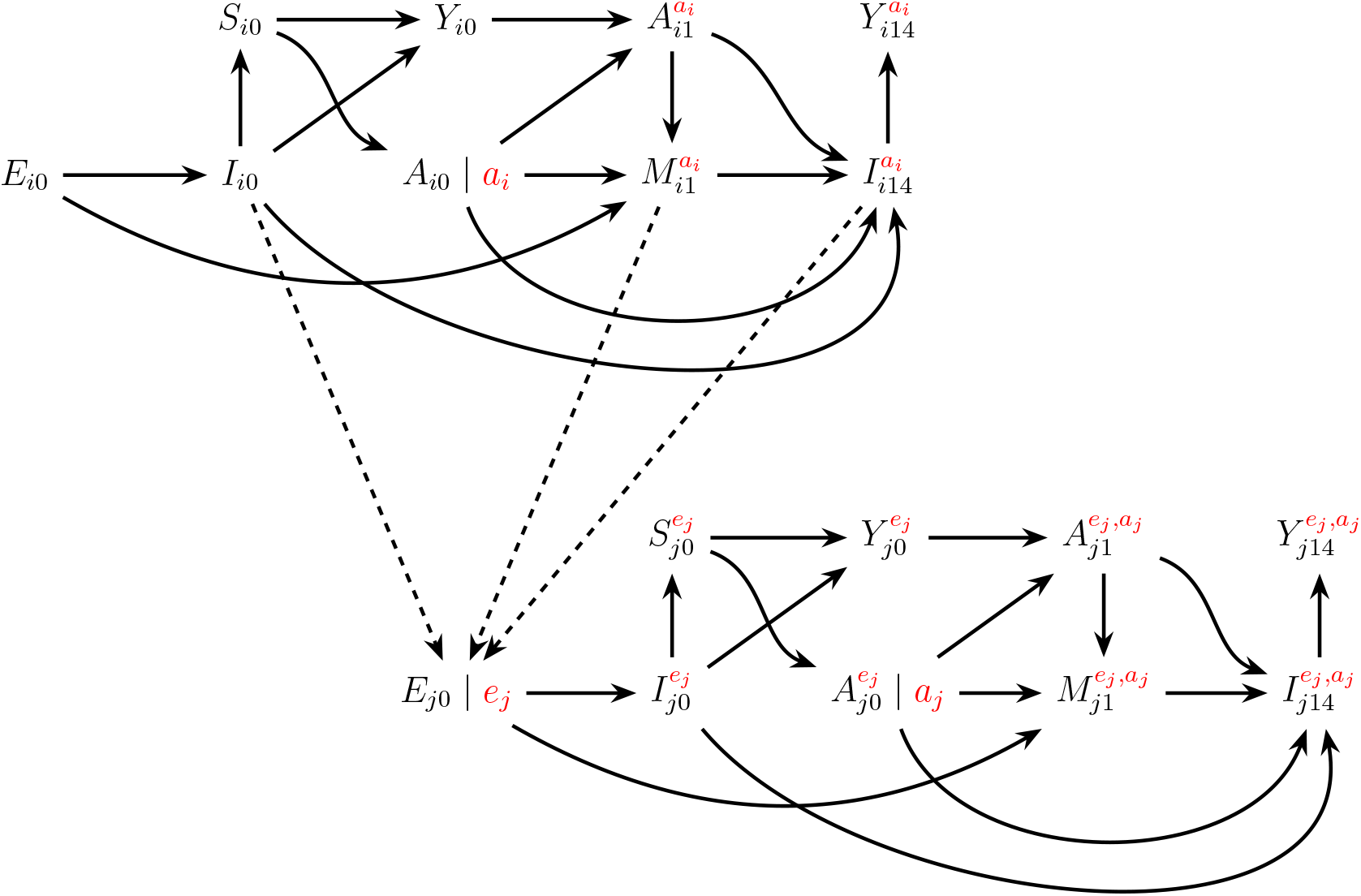
Single world intervention graph (SWIG) showing hypothetical interventions on empiric treatment of individuals *i* and *j*, and the controlled exposure on subject *j*. The dashed arrows from subject *i* to subject *j*’s exposure node *E*_*j*0_ are interrupted, reflecting an intervention that blocks indirect effects via exposure. Downstream nodes for *j* are now indexed by *e*_*j*_, not *a*_*i*_, as the exposure path has been severed by design.

Because the treatment is applied after exposure and affects subsequent exposure, the arm-specific cross-cluster comparisons considered in a mass drug administration trial (see Appendix A) do not make as much sense. Instead, we focus on the overall effect on culture results at the time infection is first suspected and a culture taken, as these have yet to be contaminated by direct treatment, but are indirectly affected by accumulating effects of treatment of previous infections, that is the potential that resistant bacteria selected by prior treatment of other patients have infected a focal patient. However, assessment of the overall effect in a post-exposure trial will depend on the sampling scheme. When all infections present to or are acquired within a health facility and the underlying source population is fully enumerated, the overall effect on resistance at time *t*, defined as contrasts such as

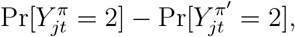

is identified under randomization by the marginal between-cluster comparison of incidence of resistant infection at admission in clusters assigned to saturation *π* versus *π*^*′*^, i.e.

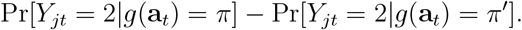

An alternative approach to estimating overall effect using odds ratio is discussed in Appendix A.

### 6.2. Observational emulation

Two-stage randomized trials are rarely conducted in practice. As before, in the absence of trial data, another option is to use observational data to attempt to emulate a two-stage trial. Emulations of two-stage trials are rarely discussed in the literature, but they are possible in theory. They would involve collecting observational data across a number of clusters with sufficient separation or buffer that the partial interference assumption holds. In this case, the observational study can be viewed as essentially a two-stage trial where allocation program **A**_*t*_ is assigned according to *f* (**A**_*t*_|**L**_*t*_, **S**_*t*_) where **L**_*t*_ are pre-treatment covariates, which is a group-level generalization of the standard conditional randomization assumption in emulations of individually-randomized trials. A consequence of this is that, unlike emulations of individually-randomized trials, emulations of two-stage trials must account for potential confounding via individual and group-level covariates. Another complication of two-stage emulations is that the group-level allocation plan that each cluster pursue may be ill-defined unless there are agreed upon treatment guidelines or policies within each cluster. Emulations require data from settings in which there is natural variation in antimicrobial strategies that were pursued across clusters. An example could be when there’s a shortage or introduction of a particular antimicrobial treatment.

## 7. Discussion

The use of causal inference methods to answer questions in acute infectious diseases where transmission plays an important role has not seen the same exponential growth as in other areas, such as the use of this methods in the study of chronic diseases [19]. Part of the difficulty in applying causal methods for ID studies stems from the fact that the assumption of no interference is often violated [20]. Additionally, most of the work in causal inference with interference has focused mainly on studies where vaccine is the primary intervention of interest.

In this paper, building on previous work in causal inference with interference, we focused on presenting how this framework could be implemented for the study of antimicrobial treatments strategies and AMR. We aimed to formalize cases of antimicrobial use employing DAGs and SWIGs to help those with interest in ID research present problems in a more unified way using tools form causal inference methods. This work continues the efforts of Ogburn and VanderWeele [14] by applying graphical representations to research problems involving interference. We also discussed how challenge trials and a two-stage randomization design (and potentially their observational emulations) could be adapted to study the indirect effects of antimicrobial treatment strategies. Traditionally, evaluations of new antimicrobial strategies have focused on assessing non-inferiority compared to the standard of care [21]. However, there is increasing interest in understanding how these strategies might affect the spread of AMR, particularly through indirect effects arising from interference between patients. In settings such as ICUs and nursing homes—where antimicrobials are extensively used among vulnerable populations— the potential for patient-to-patient transmission is significant [22, 23]. Expanding study designs to account for these indirect effects is key for a more comprehensive assessment of treatment impacts. By explicitly incorporating these effects, we can better evaluate how interventions influence both individual outcomes and the broader dynamics of resistance, making stronger stewardship recommendations.

Using DAGs and SWIGs as we have presented facilitates the clear specification of causal contrasts and assumptions. These graphical tools make the underlying causal structures explicit, allowing researchers to identify and address potential biases in studies assessing antimicrobial treatments and AMR. By clarifying the causal pathways under consideration and formalizing the nodes commonly present in ID studies, our approach enhances the evaluation of causal effect estimates in scenarios where interference is present, aligning with methodologies discussed by previous work [8, 10, 20]

The application of the methods described here are future steps that this research group plan to apply to different current clinical problems. We also hope that the present work will encourage the broader use of causal inference methods in the study of acute infectious diseases.

## Data Availability

No data was used for this paper. Code is available upon request

## Appendix A

### A.1. Representation of the Per-exposure effect in SWIGs

When estimating a per-exposure effect, we assume that all interaction with others has been eliminated, thus potential outcomes may be simplified to only depend on an individual’s intervention values. This can be seen in the SWIG in Figure A.4, and the contrast to Fig. in the main text. The intervention on *E*_*j*0_ which splits the node and blocks the indirect effects from person *i* to person *j*). Note that *Y*_*j*14_ now only depends on values assigned to subject *j*. This approach avoids the problem that in an individual RCT where interference is occurring and the only intervention is on allocation of the treatment, the intent-to-treat estimate is not guaranteed to be equal to the causal estimate [17].

### A.2.Appendix Simple 2-stage randimoization design: Mass Drug Administration (MDA)

Perhaps the most straightforward adaptation of the two-stage design for vaccines would be a design to capture the use of antibiotics irrespective of infection such as in the mass drug administration of azithromycin [24]. Originally used for treatment of trachoma, where it was judged more efficient to treat all than to test and treat only those with trachoma [25], mass azithromycin distribution to under-5 year olds in areas of high child mortality has been shown to reduce mortality risk. In this case the intervention does not condition on being previously exposed, infected, or presenting to a health facility, but rather is typically done in communitybased campaign in lower resourced settings where healthcare and drug treatments may not be readily available. Because the intervention closely resembles a vaccination campaign, very few changes are needed from the canonical two-stage vaccine design.

Under this design we can define the direct effect on resistance (i.e blood culture at 14 days after administration as positive for resistant strains) as

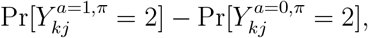

which is identified under randomization by the within-cluster comparison of those treated with antimicrobials (*A*_*kj*_ = 1) versus those untreated (*A*_*kj*_ = 0) among clusters assigned to treatment saturation *π*, i.e.

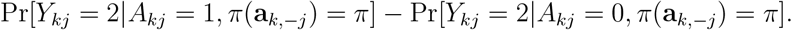

For the direct effects on other outcomes like mortality or susceptible infection we simply replace *Y*_*kj*_ with the appropriate outcome.

We can also define the indirect effects at different saturation levels *π* versus *π*^*′*^ through

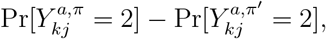

which is identified under randomization by the between-cluster comparison of clusters assigned to saturation *π* versus *π*^*′*^, i.e.

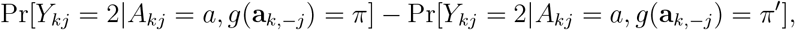

where we could consider indirect effects on antimicrobial saturation separately on those treated or on those untreated.

Finally, we can define the population-level overall effect of saturation level *π* versus *π*^*′*^ as

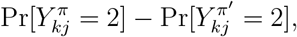

which is identified under randomization by the marginal between-cluster comparison of clusters assigned to saturation *π* versus *π*^*′*^, i.e.

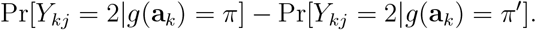

Such studies have been performed for multiple outcomes [24, 26]

### Appendix A.3. Incomplete sampling and estimation of overall effect

Alternatively, when we do not observe the outcome for all individuals in a cluster or when the sampling fractions are unknown, the assessment of the overall effect is more challenging. This may be the case for health facility based interventions where the catchment area is not fully enumerated or when both facility-based and community spread of infection or commensal carriage is possible, but only individuals with severe enough infection to seek care are observed. One work-around when all suspected infections are cultured upon admission is to use the between-cluster ratio of odds that presenting infection is susceptible or resistant to treatment as a proxy for the overall effect of treatment saturation *π* versus *π*^*′*^, i.e.

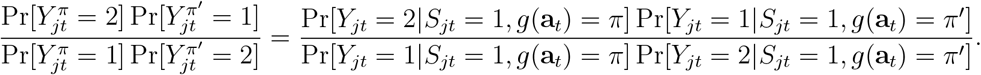

## References

1. Organization world health, w. h. Antimicrobial resistance: Report by the Director-General tech. rep. (World Health Organization, 2020).

2. Lipsitch, M. & Samore, M. H. Antimicrobial use and antimicrobial resistance: a population perspective. Emerg Infect Dis 8, 347–354. doi:10.3201/eid0804.010312 (2002).

3. Halloran, M. E. & Struchiner, C. J. Causal Inference in Infectious Diseases. Epidemiology 6, 142–151 (1995).

4. Huskins, W. C. et al. Intervention to Reduce Transmission of Resistant Bacteria in Intensive Care. New England Journal of Medicine 364, 1407–1418. doi:10.1056/NEJMoa1000373 (2011).

5. Kunkel, A., Colijn, C., Lipsitch, M. & Cohen, T. How could preventive therapy affect the prevalence of drug resistance? Causes and consequences. Philosophical Transactions of the Royal Society B: Biological Sciences 370, 20140306. doi:10.1098/rstb.2014.0306 (2015).

6. Duijn, P. J. v. et al. The effects of antibiotic cycling and mixing on antibiotic resistance in intensive care units: a cluster-randomised crossover trial. English. The Lancet Infectious Diseases 18, 401–409. doi:10.1016/S1473-3099(18)30056-2 (2018).

7. Sobel, M. E. What Do Randomized Studies of Housing Mobility Demonstrate?: Causal Inference in the Face of Interference. Journal of the American Statistical Association 101, 1398–1407 (2006).

8. Hudgens, M. G. & Halloran, M. E. Toward Causal Inference With Interference. Journal of the American Statistical Association 103, 832–842. doi:10.1198/016214508000000292 (2008).

9. Tchetgen Tchetgen, E. J. & VanderWeele, T. J. On causal inference in the presence of interference. eng. Statistical Methods in Medical Research 21, 55–75. doi:10.1177/0962280210386779 (2012).

10. VanderWeele, T. J. & Tchetgen Tchetgen, E. J. Effect partitioning under interference in two-stage randomized vaccine trials. Statistics & probability letters 81, 861–869. doi:10.1016/j.spl.2011.02.019 (2011).

11. Chanderraj, R. et al. Vancomycin-Resistant Enterococcus Acquisition in a Tertiary Care Hospital: Testing the Roles of Antibiotic Use, Proton Pump Inhibitor Use, and Colonization Pressure. Open Forum Infectious Diseases 6, ofz139. doi:10.1093/ofid/ofz139 (2019).

12. Adamu, Y., Puig-Asensio, M., Dabo, B. & Schweizer, M. L. Comparative effectiveness of daptomycin versus vancomycin among patients with methicillin-resistant Staphylococcus aureus (MRSA) bloodstream infections: A systematic literature review and meta-analysis. PLOS ONE 19, e0293423. doi:10.1371/journal.pone.0293423 (2024).

13. Heath, M. R. et al. Gut colonization with multidrug resistant organisms in the intensive care unit: a systematic review and meta-analysis. Critical Care 28, 211. doi:10.1186/s13054-024-04999-9 (2024).

14. Ogburn, E. L. & VanderWeele, T. J. Causal Diagrams for Interference. Statistical Science 29, 559–578. doi:10.1214/14-STS501 (2014).

15. Richardson T R. J. Single World Intervention Graphs (SWIGs) : A Unification of the Counterfactual and Graphical Approaches to Causality in (2013).

16. Adams-Phipps, J. et al. A Systematic Review of Human Challenge Trials, Designs, and Safety. Clinical Infectious Diseases: An Official Publication of the Infectious Diseases Society of America 76, 609–619. doi:10.1093/cid/ciac820 (2022).

17. O’Hagan, J. J., Lipsitch, M. & Hernán, M. A. Estimating the Per-Exposure Effect of Infectious Disease Interventions. en-US. Epidemiology 25, 134. doi:10.1097/EDE.0000000000000003 (2014).

18. Eyal, N. & Lipsitch, M. Testing SARS-CoV-2 vaccine efficacy through deliberate natural viral exposure. eng. Clinical Microbiology and Infection: The Official Publication of the European Society of Clinical Microbiology and Infectious Diseases 27, 372–377. doi:10.1016/j.cmi.2020.12.032 (2021).

19. Scola, G. et al. Implementation of the trial emulation approach in medical research: a scoping review. en. BMC Medical Research Methodology 23, 186. doi:10.1186/s12874-023-02000-9 (2023).

20. Halloran, M. E. & Hudgens, M. G. Causal Inference for Vaccine Effects on Infectiousness. en. The International Journal of Biostatistics 8, 1–40. doi:10.2202/1557-4679.1354 (2012).

21. Lanini, S. et al. Non-inferiority versus superiority trial design for new antibiotics in an era of high antimicrobial resistance: the case for post-marketing, adaptive randomised controlled trials. eng. The Lancet. Infectious Diseases 19, e444–e451. doi:10.1016/S1473-3099(19)30284-1 (2019).

22. Van den Dool, C., Haenen, A., Leenstra, T. & Wallinga, J. The Role of Nursing Homes in the Spread of Antimicrobial Resistance Over the Healthcare Network. Infection Control and Hospital Epidemiology 37, 761–767. doi:10.1017/ice.2016.59 (2016).

23. Wunderink, R. G. et al. Antibiotic Stewardship in the Intensive Care Unit. An Official American Thoracic Society Workshop Report in Collaboration with the AACN, CHEST, CDC, and SCCM. Annals of the American Thoracic Society 17, 531–540. doi:10.1513/AnnalsATS.202003-188ST (2020).

24. Peterson, B. et al. Assessment of Spillover of Antimicrobial Resistance to Untreated Children 7–12 Years Old After Mass Drug Administration of Azithromycin for Child Survival in Niger: A Secondary Analysis of the MORDOR Cluster-Randomized Trial. Clinical Infectious Diseases: An Official Publication of the Infectious Diseases Society of America 79, 1136–1143. doi:10.1093/cid/ciae267 (2024).

25. Mariotti, S. P. New Steps toward Eliminating Blinding Trachoma. New England Journal of Medicine 351, 2004–2007. doi:10.1056/NEJMe048205 (2004).

26. O’Brien, K. S. et al. Azithromycin distribution and childhood mortality in compliance-related subgroups in Niger: complier average causal effect and spillovers in a cluster-randomized, placebo-controlled trial. International Journal of Epidemiology 51, 1775–1784. doi:10.1093/ije/dyab198 (2022).

